# Effective Connectivity in Subcortical Visual Structures in *De Novo* Patients with Parkinson’s Disease

**DOI:** 10.1101/2021.07.01.21259832

**Authors:** Emmanuelle Bellot, Louise Kauffmann, Véronique Coizet, Sara Meoni, Elena Moro, Michel Dojat

**Affiliations:** University Grenoble Alpes, Inserm U1216, CHU Grenoble Alpes, Grenoble Institute of Neurosciences, Grenoble, France; Laboratory of Psychology and Neurocognition, CNRS UMR 5105, Grenoble, France; Movement Disorders Unit, Division of Neurology, CHU Grenoble Alpes, Grenoble, France

**Keywords:** Dynamic Causal Modelling, fMRI, Human Vision, Neuroimaging, Superior Colliculus

## Abstract

**Background:** PD is associated with non-motor symptoms appearing before the motor symptoms onset. Recent studies report dysfunctions of visual structures at early stages of PD.

**Objective:** This study addresses effective connectivity in the visual network of PD patients.

**Methods:** Using brain functional MRI and Dynamic Causal Modeling analysis, we investigated the connectivity between the superior colliculus, the lateral geniculate nucleus and the primary visual area V1 in 22 *de novo* untreated PD patients and six months after starting dopaminergic treatment compared to age-matched healthy controls.

**Results:** Our findings indicate that the superior colliculus drives cerebral activity for luminance contrast processing both in healthy controls and untreated PD patients. The same effective connectivity was observed with neuromodulatory differences in terms of neuronal dynamic interactions. The modulation induced by luminance contrast changes of the superior colliculus connectivity (self-connectivity and connectivity to the lateral geniculate nucleus) was inhibited in PD patients (effect of contrast: *p* = 0.79 and *p* = 0.77 respectively). The introduction of dopaminergic medication failed to restore the effective connectivity modulation observed in the healthy controls.

**Interpretation:** The deficits in luminance contrast processing in PD seem due to a deficiency in connectivity adjustment from the superior colliculus to the lateral geniculate nucleus and to V1. Administration of a dopaminergic treatment over six months was not able to normalize the observed alterations in inter-regional coupling. These findings highlight the presence of early dysfunctions in primary visual areas, which might be used as early markers of the disease.

## Introduction

Parkinson’s disease (PD) is a progressive neurodegenerative disorder affecting more than six million people worldwide.^1^ The onset of the pathological process is believed to start 10-20 years before the classical motor manifestations (tremor, rigidity and bradykinesia), the so-called prodromal phase of PD.^2^ Several non-motor symptoms^3^ especially sensory dysfunctions like hyposmia^4,5^ and oculo-visual affections in pupil reactivity, color vision or visuo-motor adaption^6^, develop during this prolonged prodromal phase. With the progress of the disease, several dysfunctions along the visual pathway appear in the early stages of PD^7^, from the retina^8^ to higher visual brain areas, starting from V1 to the fusiform gyrus.^9^ PD patients show visuospatial and visuoperceptual deficits, which reflect structural changes in temporoparietal cortical regions.^10^ Visual hallucinations are reported to be common in advanced PD.^11^ Because neuron degeneration starts in subcortical structures before invading the cortex, alterations in the first steps of visual processing as the superior colliculus (SC) and the lateral geniculate nucleus (LGN) are expected and may constitute early biomarkers of the disease.

Neuroimaging is a powerful tool for improving our understanding of both regional specific dysfunction^12^ and brain connectivity changes in PD.^13^ Several studies have explored resting-state functional connectivity (i.e. a statistical dependency between the activity of regions)^14-18^. Fewer studies examined changes in effective connectivity, i.e. how the network of activity between brain regions is affected by the pathology, focusing on the detection of changes in cortical motor network coupling^19-25^ and modulatory actions of deep brain stimulation.^26,27^ No study has concerned the subcortical and cortical effective connectivity in the visual network and how it is affected by PD.

Using brain functional MRI (fMRI) in healthy subjects of different ages, we showed an increase in the Blood Oxygen Level Dependent (BOLD) responses along the visual pathway within the SC, the LGN and V1 in response to increasing luminance contrast in the range of 1-9%, and an alteration with normal aging (see Supplementary Material (SM), Figure 1 for a scheme of the fMRI experience).^28^ In a subsequent study in *de novo* PD patients, we found already present a functional deficit in both the SC and the LGN that was not compensated by dopaminergic treatment at six months (see SM: Figure 2).^29^ The activation in our three regions of interest (ROIs) was correlated with luminance contrast modulation but did not inform us about the ROIs interactions. The main aim of the present study was to further investigate these findings by addressing brain connectivity, i.e. interactions between our visual ROIs, and exploring how visual information processing in this network might be already affected in *de novo* untreated PD patients. We hypothesize that the effective connectivity between the SC, the LGN and V1 would change in *de novo* PD patients compared to age-matched healthy controls and that the initiation of dopaminergic treatment over six months would not restore the inter-regional coupling. We analyzed effective connectivity using Dynamic Causal Modelling (DCM), a probabilistic (Bayesian) modelling technique that estimates the coupling between selected brain regions based on fMRI data i.e., their BOLD responses to a specific stimulation, in our case visual stimuli.

**Figure 1.**
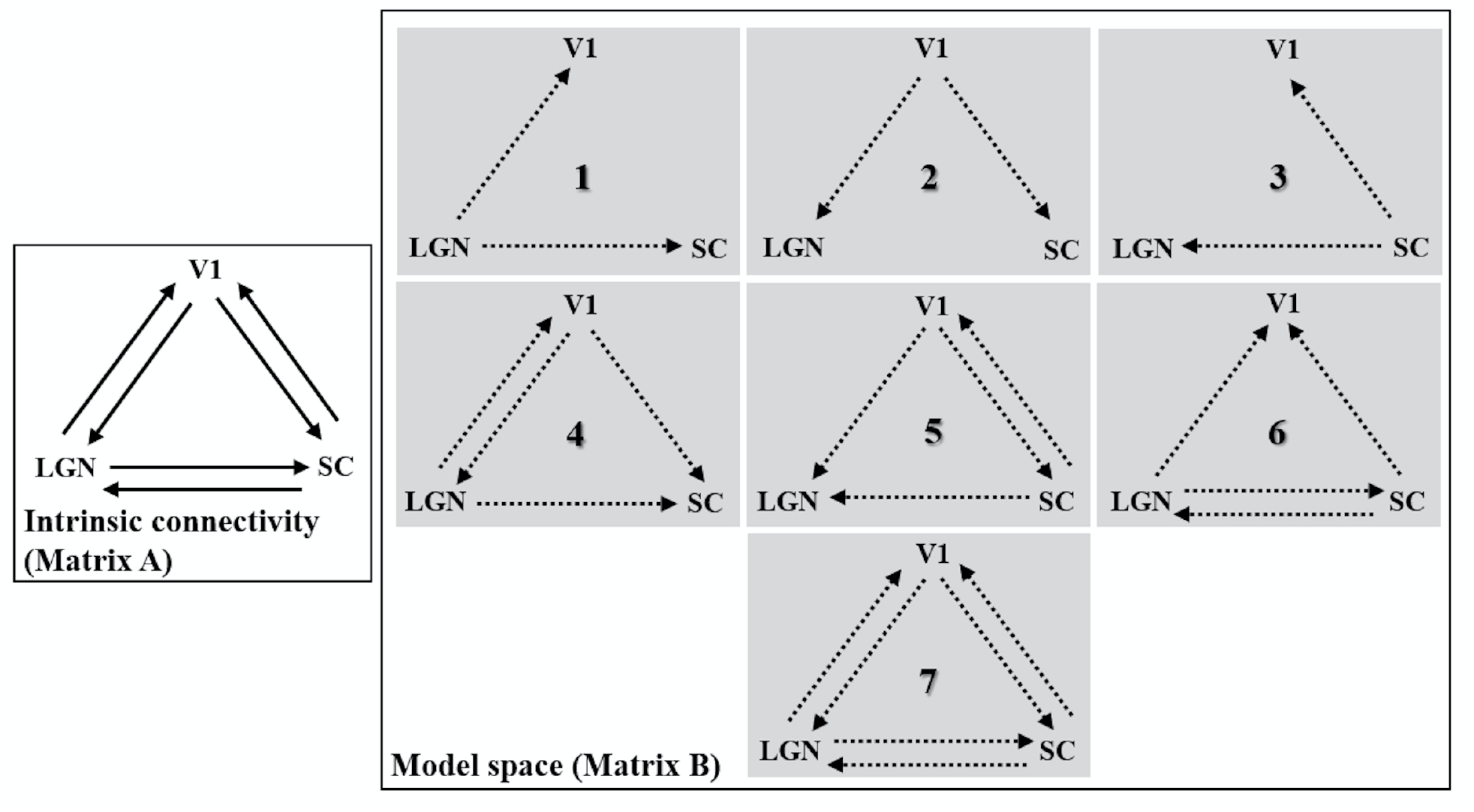
DCM model space of the modulations of effective connectivity between our regions of interest. The intrinsic connectivity (Matrix A) was taken to be complete (bidirectional) between regions. Activations in responses to the first luminance level (1% luminance contrast) were used as input in LGN and SC (matrix C) while activations to luminance contrast (3, 5 and 9%) could modulated effective connectivity (matrix B) depending on each model. Each model included unilateral modulation between one region to the two others (M1-M3). Additionally, there could be bilateral modulation from V1 to LGN or to SC (M4 and M5) or between LGN and SC (M6). Finally, all connections could be modulated (M7). This yields to seven models. Additionally, each model could include self-modulation in SC leading to 14 models. SC = superior colliculus, LGN = lateral geniculate nucleus, V1 = primary visual area. Solid arrows: intrinsic connections. Dashed arrows: connections modulated by luminance.

**Figure 2.**
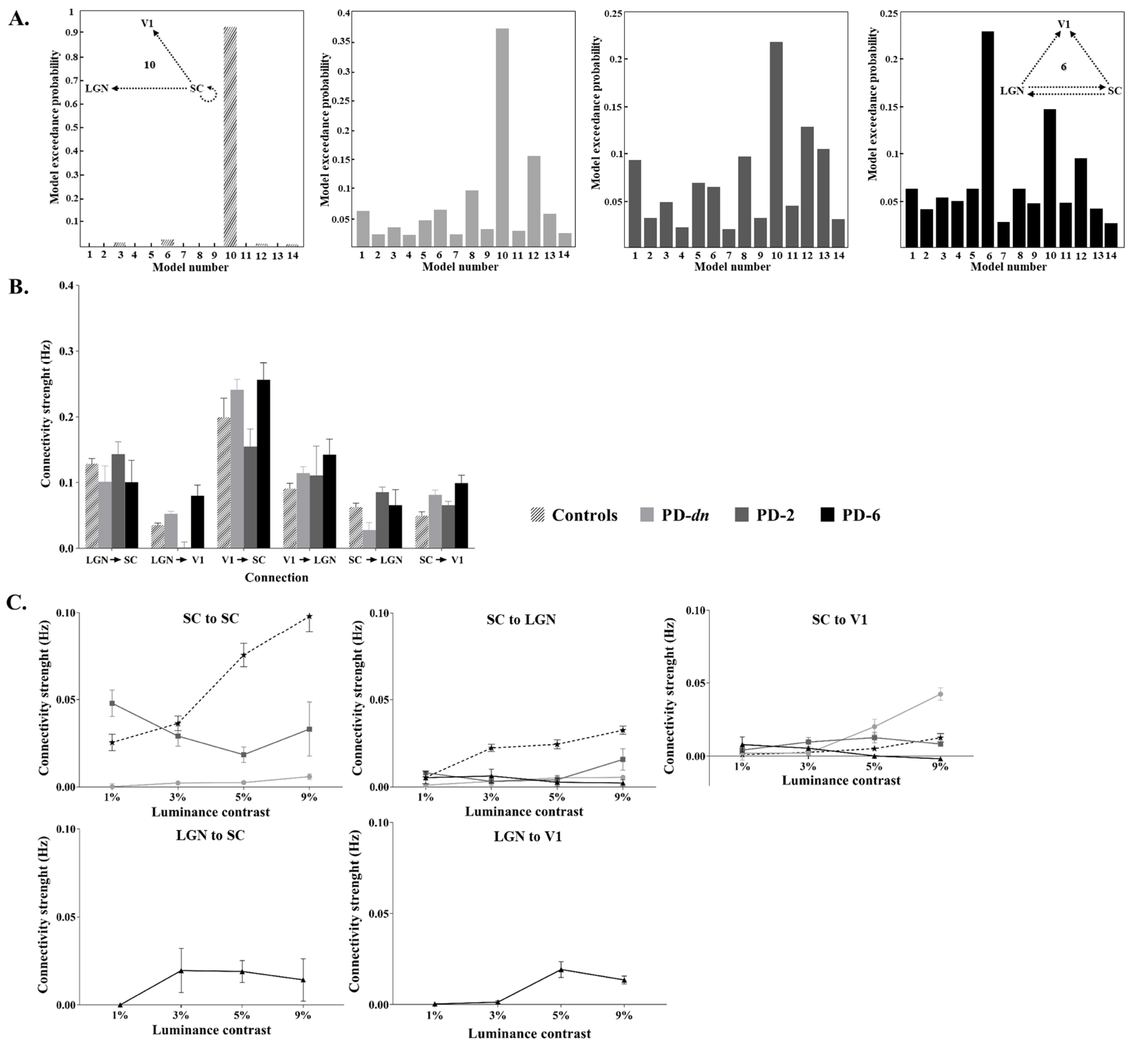
Effective connectivity in control and patients. A. Exceedance probabilities of the models: from left to right: matched controls (CT-22); *de novo* PD patients (PD-*dn*); PD patients 2-months after treatment (PD-*2*); PD patients 6-months after treatment (PD-*6*). Model 10 (see Insert) was the best model with random effect Bayesian model selection (RFX) for CT-22, PD-*dn* and PD-*2*, whereas model 6 (see Insert) was the best model for PD-*6*. B. Intrinsic connections parameters of the selected model for PD patients *vs*. matched controls. C. Intrinsic connection modulation with luminance contrast. Dashed line and stars: CT-22. Bold line: PD patients; Circle: PD-dn, Square: PD-2, Triangle: PD-6. Intrinsic connection and modulatory parameters are expressed in Hz. Same abbreviations than in Figure 2. The vertical bars represent standard errors.

## Materials and methods

### Participants

Thirty healthy subjects (CT-30) and 22 *de novo* PD patients were recruited at the Movement Disorders Center of the Centre Hospitalier Universitaire of Grenoble (Grenoble, France). Inclusion criteria were: recent diagnoses of PD (< 1year of motor symptoms’ onset) according to the Movement Disorder Society clinical diagnostic criteria of Parkinson’s disease (MDS)^30^, stage 1 or 2 of the Hoehn and Yahr scale^31^, with no psychiatric disorders; no antiparkinsonian treatment. A subset of the Control’s sample, CT-22, was age-matched with PD patients. Additionally, eight patients from the PD-*dn* group, who had started L-dopa or dopamine agonist treatment after the first brain MRI study, were further evaluated two times: at six weeks to two months (session S1, resp. PD-*2*), and at six months (session S2, resp. PD-*6*) after anti-PD treatment onset. All the selected participants were enrolled in our two previous studies.^28, 29^ All participants provided written informed consent to the study. The study was approved by the local ethical committee (Comité de Protection des Personnes Sud-Est V, ID RCB-2012-A00310-43 and ID RCB-2014-A01835-42) and registered with ClinicalTrial.gov (NCT02488395).

### fMRI design and procedure

Both visual stimuli and MRI acquisition were previously described in details.^28^ Briefly, subjects were presented in each visual hemi-field with a series of achromatic checkerboards, viewed via a mirror fixed on the head coil and flashing at a frequency of 4Hz on a grey background, with four levels of luminance contrast (1, 3, 5 and 9%) to avoid the SC saturation.^32^ We used a block-design visual paradigm with each luminance contrast level presented in separate 12-s blocks. For an accurate activation localization in SC and limit partial volume effect, we used at 3 T a 32-channel SENSE coil, a spatial resolution of 1.5 mm in each direction for EPI sequence, with an acquisition volume centered on the SC structure. A high-resolution T1-weighted (MPRAGE) sequence and a T1-weighted FGATIR sequence were acquired to facilitate the manual delineation of SC and LGN (see details in SM). The cardiac signal was indirectly recorded at 100 Hz using a finger photoplethysmography. Functional data analysis was performed using the single-participant general linear model (GLM) for block-designs with SPM12 (Wellcome Department of Imaging Neuroscience, London, U.K.) implemented in Matlab. The design matrix was constructed in order to remove possible motion components and cardio-respiratory effects, the main cause of source of noise in the BOLD signal in the SC (see SM).

### Statistical analysis

#### Dynamic causal modeling for BOLD responses

DCM^33,34^ explores changes in the effective connectivity, i.e. how the influence exerted by one region over other regions can be affected by the experimental conditions. DCM allows to make inferences about the neural mechanisms underlying the observations, here the BOLD time course in our three ROIs (see SM for Time series extraction). Causal network models representing our hypotheses about these mechanisms were constructed and their parameters estimated (see SM about the specification of such models in DCM). Three sets of parameters are considered: the driving input parameters (e.g., visual stimulation), the intrinsic connection parameters and the modulatory parameters that define how the effective connectivity may be modulated by, in our case, luminance contrast variations. All the intrinsic connections are not necessarily affected by the modulation leading to alternative plausible models that should be estimated.

Our aim was to assess whether differences observed in fMRI responses in PD patients compared to age-matched controls, could be explained by the fact that PD and dopaminergic treatment change the architecture of effective connectivity between the visual ROIs, and/or change the connectivity strength and its capability to be modulated by luminance contrast. Thus, we first tested for each group which model best explained our data, second whether the winning model differed according to the group, then revealing a change in connectivity architecture. Finally, we examined and compared across groups the estimated parameters of their respective winning model.

#### Model space

We considered the six possible endogenous connections between our three structures (see Figure 1, matrix A of the network). Neurophysiological studies in primates have shown bidirectional structural connections between the LGN and the V1^35-37^ as well as structural connections from the SC to the LGN.^38-40^ Connections from the V1 to the SC have also been identified in macaque monkeys.^41-45^ Indeed, injections of anterograde tracers in non-human primates indicates that the SC receives input from most areas of visual cortex.^46^ However, to our knowledge, no direct inputs from the SC to the V1 and the LGN to the SC have been reported so far. It should be noted that effective connectivity between two regions does not necessarily imply direct anatomical connections as the influence of one region over another can be mediated by other relay structures. For example, the SC and the V1 are connected via the Pulvinar.^47^ Our visual stimuli entered all models as a driving input to the two subcortical nodes, the LGN and the SC, both receiving direct visual inputs from the retina.

#### Connectivity Modulation

We designed a model space by varying the subset of connections which were modulated by luminance contrast variations. Our previous fMRI results showed that luminance contrast fluctuations modulated the activations in the visual regions of interest (increased response with increasing contrast, see SM: Figure 2). For the effective connectivity study our rational was that the lowest contrast condition (1%) constituted a baseline from which the response was further modulated by contrast increase. We therefore used the remaining contrast conditions (3%, 5% and 9%) as modulating inputs, to explore the connectivity modulation with increasing contrasts relative to the 1% contrast condition. These transient perturbations were implemented as bilinear terms. We considered a bilinear evolution based on our previous results that showed a linear relation between BOLD signal and luminance contrast in our three regions in the variation range used.^28^ We considered a series of seven models for testing the modulatory influence of luminance contrast on effective connectivity from the LGN, V1 or the SC, to the two other structures (V1 and SC, LGN and SC or LGN and V1, cases 1 to 3 respectively), or from V1 + LGN, V1 + SC or LGN + SC on the SC, LGN or V1, respectively (cases 4 to 6 respectively), as well as all the possible modulations (case 7). We also added models with the same structure and modulations as models 1 to 7 but with an additional self-modulation of the SC leading to a total of fourteen models to compare (see Figure 1). These models (8 to 14) were used to account for the possibility of a dysfunction of the SC self-modulation in response to luminance contrast in PD. All 14 models were fed successively with the functional datasets from four groups: control subjects and untreated PD patients or treated PD patients with dopaminergic treatment (2- or 6-months uptake). Model selection and inference on parameters of the best model were made (see SM).

## Data availability

MR data supporting the results of this study are available from the corresponding author, on a collaborative basis.

## Results

### Demography

The details of the clinical characteristics of PD patients appear in Table 1. There were no significant differences between CT-22 (9 women, age 55.5 y. ± 9.4) and PD-*dn* patients (5 women, age 57.3 y. ± 10.5), in term of age (two-sample t-test; p=0.57) and gender (p=0.2).

**Table 1.** Clinical characteristics of PD patients. (this table is similar to Table 1 in^29^)

| PATIENT | SEX | AGE, yr | H&Y | Year of<br>Diagnosis | Disease<br>Onset<br>(Body side) | UPDRS | Treatment at 2 and 6 mo |
| --- | --- | --- | --- | --- | --- | --- | --- |
| 1 | M | [65-70[ | 1 | 2016 | LEFT | 18 | NA |
| 2 | F | [45-50[ | 1 | 2017 | LEFT | 11 | L-dopa/benserazide |
| 3 | M | [60-65[ | 1 | 2017 | LEFT | 31 | NA |
| 4 | M | [55-60[ | 1 | 2016 | LEFT | 11 | Pramipexole |
| 5 | M | [65-70[ | 1 | 2016 | RIGHT | 5 | NA |
| 6 | M | [75-80[ | 1 | 2016 | LEFT | 17 | NA |
| 7 | M | [55-60[ | 1 | 2015 | LEFT | 23 | Rasagiline/pramipexole |
| 8 | M | [55-60[ | 1 | 2016 | RIGHT | 15 | Rasagiline/pramipexole |
| 9 | F | [45-50[ | 1 | 2014 | LEFT | 15 | Rasagiline/L-dopa<br>benserazide |
| 10 | M | [35-40[ | 1 | 2016 | LEFT | 9 | NA |
| 11 | M | [45-50[ | 2 | 2016 | RIGHT | 11 | NA |
| 12 | F | [65-70[ | 2 | 2014 | RIGHT | 40 | Rasagiline/L-dopa<br>benserazide |
| 13 | M | [70-75[ | 2 | 2017 | LEFT | 23 | NA |
| 14 | M | [45-50[ | 2 | 2017 | RIGHT | 20 | NA |
| 15 | M | [65-70[ | 2 | 2017 | RIGHT | 25 | NA |
| 16 | M | [50-55[ | 2 | 2015 | RIGHT | 28 | Levodopa/trihyphenidyl |
| 17 | M | [50-55[ | 2 | 2016 | LEFT | 21 | NA |
| 18 | F | [55-60[ | 2 | 2016 | LEFT | 24 | NA |
| 19 | M | [65-70[ | 2 | 2015 | RIGHT | 14 | NA |
| 20 | M | [55-60[ | 2 | 2014 | RIGHT | 18 | Rasagiline/ropinirole |
| 21 | M | [40-45[ | 2 | 2016 | RIGHT | 27 | NA |
| 22 | F | [55-60[ | 2 | 2016 | RIGHT | 21 | NA |

F = Female; M = male; H&Y: Hoehn and Yahr; NA = not applicable; PD = Parkinson disease; UPDRS = Unified PD Rating Scale.

### Effective connectivity: healthy controls and PD patients

#### The best effective connectivity model for controls

In the control group (CT-22), the exceedance probabilities (xp) for the 14 models are shown in Figure 2A. As can be seen on this Figure, Model 10, which emphasizes the role of the SC, outperformed all competing models (xp=0.93). It was characterized by self-modulation by luminance in the SC and unilateral modulation from the SC to the LGN and the V1. The fitting quality of this model was excellent (see SM: Figure 3). Its parameters are reported in SM: Table 1. Additional results on the effect of age on the intrinsic connectivity for healthy controls (CT-30) are provided in SM (see SM: Figure 4 and Table 2).

#### The best effective connectivity model for PD patients

Similarly to controls, for *de novo* (untreated) PD patients (PD-*dn*), the winning model was clearly the model 10 (see Figure 2A, xp = 0.37). To note, the exceedance probability was lowered in patients (xp=0.37) compared to controls (xp ≥ 0.70) for the best candidate model because the BOLD signal variability was greater for patients. Significance of the different parameters of the model is reported in SM: Table 1.

#### Effective connectivity in PD

Since the winning model was similar in PD and Controls, suggesting that PD did not impact the architecture of connectivity between our ROIs, we then examined the connectivity parameters on this model for both groups. As for controls, all intrinsic connectivity parameters were positive, i.e. an increase in activity in each region resulted in an increase in each target region (see Figure 2B). *PD-dn* patients’ intrinsic connectivity parameters did not significantly differ from controls, as confirmed by the 2×6 ANOVA with Group (Controls CT-22, age-matched with PD patients-*vs. PD-dn* patients) as between subjects factor and Connection as within-subjects factor, revealing a main effect of Connection (*F*(5,210) = 5.82, *p* < 10^−4^, η_p_^2^ = 0.23) but no main effect of Group (*p* = 0.72) nor interaction between Group and Connection (*p* = 0.89).

#### Effect of luminance contrast modulation on the effective connectivity

A main effect of PD was observed on the SC self-modulation (*F*(1,42) = 14.03, *p* < 10^−3^, η_p_^2^ = 0.40) and on the modulation of the SC to LGN connection (*F*(1,42) = 8.46, *p* < 10^−2^, η_p_^2^ = 0.29) but not on the modulation of the SC to V1 connection (*p* = 0.15). Moreover, a main effect of luminance contrast was revealed for these three intrinsic connections (SC↔SC: *F*(3,126) = 8.08, *p* < 10^−4^, η_p_^2^ = 0.28; SC→LGN: *F*(3,126) = 3.78, *p* < 10^−2^, ηp^2^ = 0.15; SC→V1: *F*(3,126) = 6.07, *p* < 10^−3^, η_p_^2^ = 0.22). Interaction between Group and Contrast factors was only significant for the self-modulation of the SC (*F*(3,126) = 6.28, *p* < 10^−3^, η_p_^2^ = 0.23; SC→LGN: *p* = 0.14; SC→V1: *p* = 0.11) (2×4 ANOVA with Group, i.e. Controls *vs. PD-dn* patients, as between-subjects factor and Contrast as within-subjects factor, realized for each connection). To further investigate the effect of the modulation of the connections in PD, an ANOVA with Contrast as repeated-measure was realized for each connection. These analyses revealed that compared to controls, the luminance contrast modulation had a weak effect on the SC connections in *PD-dn* patients compared to controls (see Figure 2C). Indeed, the pathology hampered the self-modulation of SC (effect of contrast: *p* = 0.79) and the modulation of the SC to LGN connection (effect of contrast: *p* = 0.77). Only the SC to V1 connection was significantly modulated by luminance contrast (*F*(3,63) = 3.66, *p* = 0.02, η_p_^2^ = 0.29) in *PD-dn* patients (see Figure 2C).

### Effective connectivity in PD: Medication effects

#### The best effective connectivity model

After two months of dopaminergic treatment (PD*-2*), the same model (Model 10) was selected (xp = 0.22) (see Figure 2A). However, a different model, Model 6, was selected for PD patients after six months of dopaminergic treatment (PD*-6)* (xp = 0.23) (see Figure 2A right). The SC role is less central in model 6 compared to model 10. This suggests that after six months of treatment, the SC response is reduced. At the reverse, the LGN implication is increased with additional connections from the LGN to the V1 and to the SC. Significance of the different parameters of these models (6 and 10) is reported in SM: Table 1.

#### Effect of the dopaminergic treatment on the effective connectivity

At two and six months of treatment, intrinsic connectivity parameters from both models 6 and 10 were positive (see Figure 2B). To test the effect of dopaminergic treatment on these parameters, a 3×6 ANOVA with Group (PD*-dn vs*. PD*-2 vs*. PD*-6*) as between-subjects factor and Connection as within-subjects factor was realized. A main effect of Connection (*F*(5,175) = 6.56, *p* = 10^−4^, η_p_^2^ = 0.25) was revealed but no main effect of the Group (i.e. treatment) (*p* = 0.43) nor interaction between Group and Connection (*p* = 0.75) was observed (see Figure 2B).

#### Effects of the dopaminergic treatment on the modulation of the effective connectivity by luminance contrast

At two months of treatment a main effect of the medication was only observed on the modulation by luminance contrast of the connection SC↔SC (*F*(1,28) = 6.87, *p* = 0.02, η_p_^2^ = 0.31; SC→LGN: *p* = 0.48; SC→V1: *p* = 0.46). No main effect of Contrast was revealed for the three intrinsic connections (SC↔SC: *p* = 0.69; SC→LGN: *p* = 0.43; SC→V1: *p* = 0.07) and no interaction between Group and Contrast was observed (SC↔SC: *p* = 0.59; SC→LGN: *p* = 0.59; SC→V1: *p* = 0.12) (2×4 ANOVA with Group, i.e. PD*-dn vs*. PD*-2*, as between-subjects factor and Contrast as within-subject factor, realized for each connection) (see Figure 2C).

After six months of treatment, as characterized by the selected model (6), four connections seemed modulated by luminance contrast, i.e. SC→LGN, SC→V1, LGN→SC and LGN→V1. A 2×4 ANOVA was realized for each of the two common connections of both models 6 and 10 (SC→LGN and SC→V1) with Group (PD*-dn vs*. PD*-6*) as between-subjects factor and Contrast as within-subjects factor. No main effect of Group (SC→LGN: *p* = 0.91; SC→V1: *p* = 0.24) nor Contrast (SC→LGN: *p* = 0.98; SC→V1: *p* = 0.32) was observed on these connections. Analysis only showed an interaction between Group and Contrast for connection SC→V1 (*F*(3,84) = 2.87, *p* = 0.04, η_p_^2^ = 0.17; SC→LGN: <u>p</u> = 0.80), showing a significant difference between the two groups for 9% of contrast (*p* = 0.01) (see Figure 2C).

## Discussion

This is the first study addressing the effective connectivity in the visual network of *de novo* PD patients. First, by testing competing models of connectivity, we show that the SC drives cerebral activity in the visual network for luminance contrast processing, both in healthy controls and PD patients off medication. Second, our results demonstrate that the corresponding deficit in contrast processing reported early in PD patients seems related to a deficiency in connectivity adjustment from the SC to the LGN and the primary visual area V1. Dopaminergic treatment over six months is not sufficient to normalize this abnormal connectivity.

### Pivotal role of the SC for luminance contrast processing

For healthy subjects, the DCM analysis supports a model in which luminance contrast modulated the SC self-connectivity, as well as effective connectivity from the SC to the V1 and to the LGN. For these subjects, an increase in luminance contrast resulted in an increase of the SC self-connectivity for all age ranges and for the SC to LGN and V1 couplings except for elderlies (see SM: Figure 4C and SM: Table 2). Our data also indicate that connectivity strength within and from the SC decreases with normal aging (see SM: Figure 4B and SM: Table 2). This decrease may explain the age-related luminance contrast sensitivity loss observed in these regions with BOLD measurements and also revealed based on behavioral responses.^28^ Due to the small size of our regions of interest, and in order to maximize the signal to noise ratio, we pooled the right and left hemisphere data for each ROI for each subject. Indeed, for the SC that plays a central role in our study, visual information from the right to the left colliculus can be transferred via the inter-collicular commissure or other inter-hemispheric tracks.^48^

### Effect of PD on effective connectivity

We addressed nonmotor impact of PD and studied effective connectivity of the visual system measured by DCM. DCM relies on fMRI measures. In fMRI, the measured BOLD signal reflects changes in hemodynamic parameters induced by neural activation and is considered as an index of cerebral activity. We have previously shown that the observed BOLD signal variations were due to neural activity changes and were not a consequence of perfusion alterations both in healthy subjects^28^ and PD patients.^29^ The present study has investigated the alterations due to PD of the first steps of the visual processing at the cortical level in the V1 and at the subcortical level in the LGN and the SC. The SC is a complex sensory-motor brainstem structure that acts as a sentinel for detecting sudden environmental changes and responding to these changes. It integrates multimodal sensory information from visual, auditory, and tactile sources; generates outputs for gaze, head, and arm movement; and sends priority signals to the *substantia nigra pars compacta* and the intralaminar nucleus of the thalamus. The SC abnormal responses to visual stimuli have been demonstrated in a PD rat model^49^ and more recently in humans.^29^ In a post-mortem study,^50^ neurodegenerative changes in the SC tissue were found in Lewy bodies dementia that may contribute to visual attention deficits and visual hallucinations; manifestations which are also present in PD patients.^11,51^

Our DCM analysis revealed that the best model explaining the data in terms of intrinsic and modulated connectivity was the same for healthy controls and *de novo* untreated PD patients (see respectively Figure 2A). As previously underlined, this model emphasized the pivotal role of the SC for luminance contrast processing. However, we found that the modulation of the SC connectivity (self-connectivity and connectivity to the LGN) by luminance contrast was inhibited in PD patients (see SM: Table 1). This suggests that while PD patients have the same effective connectivity as healthy controls the neuromodulation is different in terms of neuronal dynamic interactions. The SC-V1 connection modulation by luminance contrast was preserved at this stage of PD, thus explaining why PD patients did not report about visual deficits.

### Effect of PD treatment

For brain regions involved in manual movement, some studies^22,23^ showed that dopaminergic medication normalized to some extent effective connectivity in PD patients. Similarly, a relative normalization of visuo-parietal connectivity in PD patients with dopaminergic medication during cued and uncued writing was found.^52^ Here, the introduction of the dopaminergic treatment during 2 or 6 months did not permit to recover from abnormal SC connectivity to other brain regions. However, our analyses suggest that after six months of anti-PD treatment, the LGN plays a more important role (see model 6, Figure 2A) in the investigated network although the effective connectivity modulation by luminance contrast was not recovered (see Figure 2C). It has been suggested^49^ that the alteration of the SC response may occur as a compensatory mechanism in response to the inhibition exerted by the loss of dopaminergic neurons in the substantia nigra reticulata. Therefore, dopaminergic treatment may not be adapted to rapidly (here 6 months uptake) reverse this process.^23^

Our study has several limitations. Although the reasonable power for DCM analysis was reached,^53^ the present study remains limited by the small size of the subject samples. Indeed, to assess the effect of medication on the inter-regional coupling, only half of the initial patient’s sample was under medication. Our results need to be replicated on a larger population. Furthermore, we did not consider a hemispheric difference in effective connectivity that could be induced by the lateralization of the disease. Indirect neuronal information provided by the BOLD signal allows defining connectivity models at a system level using DCM but hampers the interpretation of effective connectivity in terms of inhibition or excitation at the neuronal level. The poor temporal resolution of BOLD response did not allow to temporal dynamics of the neuronal model. Based on the literature and our previous results both in Human and PD animal model, we consider that the colliculus is a key sensory structure early impacted by PD. Recently, we have demonstrated that its superior part, involved in visual information processing, was early altered in its functionality by PD. Our main goal here was to investigate the role of SC in regional coupling both in normal and pathological conditions. Then, we introduced models with or without SC self-modulation. Introduction of more models, for instance with self-modulation on the other nodes, could be explored at the expense of the robustness of the results obtained. Finally, because DCM analyses becomes less effective as the number of regions of interest increases, we restricted our hypothesis testing to a small number of nodes, namely SC, LGN and V1, which were the most informative. It should be noted that the missing regions do not affect the estimates in the regions of interest because influences from outside the restricted network are modelled with endogenous fluctuations.^54,55^ Nevertheless, we left several other activated regions unexplored. For example, neuroanatomical studies suggest that the SC forms a subcortical path that bypasses the striate cortex V1 and projects to the amygdala via the pulvinar. Therefore, further research concerning the impact of the SC alterations on connectivity with these structures may help to precise the effects of the disease.

In summary, these findings provide useful insights concerning brain alterations occurring early in PD patients, and highlights the presence of early dysfunctions in primary visual areas, which might be used as early markers of the disease.

## Data Availability

All the data are available on request for scientific research objectives (contact M. Dojat). The data are not publicly available due to privacy restrictions, stated in the document approved by the Institutional Review Board.

## Acknowlegements

The authors are thankful for the patients and healthy controls who took part in this study.

## Author Roles

(1) Research project: A. conception, B. organization, C. execution; (2) statistical analysis: A. design, B. execution, C. review and critique; (3) manuscript: A. writing of the first draft, B. review and critique.

E.B.: 1B, 1C, 2A, 2B, 2C, 3A.

L.K.: 2A, 2C, 3B.

V.C.: 1A, 1B.

S.M.: 1B, 1C, 3B.

E.M.: 1A, 1B, 3B.

M.D.: 1A, 1B, 1C, 2A, 2C, 3B.

## Abbreviations

BOLD: Bold Oxygen Level Dependent
DCM: Dynamic Causal Modelling
fMRI: functional Magnetic Resonance Imaging
LGN: Lateral Geniculate Nucleus
PD: Parkinson’s Disease
ROI: Region Of Interest
SC: Superior Colliculus
V1: primary visual cortex.

## References

1. Collaborators GPsD. Global, regional, and national burden of Parkinson’s disease, 1990-2016: a systematic analysis for the Global Burden of Disease Study 2016. ZLancet Neurol. 2018;17(11):939–953.

2. Mahlknecht P, Seppi K, Poewe W. The Concept of Prodromal Parkinson’s Disease. J Parkinsons Dis. 2015;5(4):681–697.

3. Siderowf A, Lang AE. Premotor Parkinson’s disease: concepts and definitions. Mov Disord. 2012;27(5):608–616.

4. Lerner A, Bagic A. Olfactory pathogenesis of idiopathic Parkinson disease revisited. Mov Disord. 2008;23(8):1076–1084.

5. Siderowf A, Jennings D, Eberly S, et al. Impaired olfaction and other prodromal features in the Parkinson At-Risk Syndrome Study. Mov Disord. 2012;27(3):406–412.

6. Armstrong RA. Oculo-Visual Dysfunction in Parkinson’s Disease. J Parkinsons Dis. 2015;5(4):715–726.

7. Weil RS, Schrag AE, Warren JD, Crutch SJ, Lees AJ, Morris HR. Visual dysfunction in Parkinson’s disease. Brain. 2016;139(11):2827–2843.

8. Ahn J, Lee JY, Kim TW, et al. Retinal thinning associates with nigral dopaminergic loss in de novo Parkinson disease. Neurology. 2018;91(11):e1003–e1012.

9. Cardoso EF, Fregni F, Maia FM, et al. Abnormal visual activation in Parkinson’s disease patients. Mov Disord. 2010;25(11):1590–1596.

10. Pereira JB, Junque C, Marti MJ, Ramirez-Ruiz B, Bargallo N, Tolosa E. Neuroanatomical substrate of visuospatial and visuoperceptual impairment in Parkinson’s disease. Mov Disord. 2009;24(8):1193–1199.

11. Diederich NJ, Fenelon G, Stebbins G, Goetz CG. Hallucinations in Parkinson disease. Nat Rev Neurol. 2009;5(6):331–342.

12. Grafton ST, Woods RP, Tyszka M. Functional imaging of procedural motor learning: Relating cerebral blood flow with individual subject performance. Hum Brain Mapp. 1994;1(3):221–234.

13. Rowe JB. Connectivity analysis is essential to understand neurological disorders. Front Syst Neurosci. 2010;4.

14. Hacker CD, Perlmutter JS, Criswell SR, Ances BM, Snyder AZ. Resting state functional connectivity of the striatum in Parkinson’s disease. Brain. 2012;135(Pt 12):3699–3711.

15. Helmich RC, Derikx LC, Bakker M, Scheeringa R, Bloem BR, Toni I. Spatial remapping of cortico-striatal connectivity in Parkinson’s disease. Cereb Cortex. 2010;20(5):1175–1186.

16. Skidmore F, Korenkevych D, Liu Y, He G, Bullmore E, Pardalos PM. Connectivity brain networks based on wavelet correlation analysis in Parkinson fMRI data. Neurosci Lett. 2011;499(1):47–51.

17. van Eimeren T, Monchi O, Ballanger B, Strafella AP. Dysfunction of the default mode network in Parkinson disease: a functional magnetic resonance imaging study. Arch Neurol. 2009;66(7):877–883.

18. Wu T, Wang L, Chen Y, Zhao C, Li K, Chan P. Changes of functional connectivity of the motor network in the resting state in Parkinson’s disease. Neurosci Lett. 2009;460(1):6–10.

19. Buijink AWG, Van Der Stouwe AMM, Broersma M, et al. Motor network disruption in essential tremor: a functional and effective connectivity study. Brain. 2015;138(10):2934–2947.

20. Dirkx MF, den Ouden H, Aarts E, et al. The Cerebral Network of Parkinson’s Tremor: An Effective Connectivity fMRI Study. J Neurosci. 2016;36(19):5362–5372.

21. Marreiros AC, Cagnan H, Moran RJ, Friston KJ, Brown P. Basal ganglia-cortical interactions in Parkinsonian patients. Neuroimage. 2013;66:301–310.

22. Nettersheim FS, Loehrer PA, Weber I, et al. Dopamine substitution alters effective connectivity of cortical prefrontal, premotor, and motor regions during complex bimanual finger movements in Parkinson’s disease. Neuroimage. 2019;190:118–132.

23. Palmer SJ, Eigenraam L, Hoque T, McCaig RG, Troiano A, McKeown MJ. Levodopa-sensitive, dynamic changes in effective connectivity during simultaneous movements in Parkinson’s disease. Neuroscience. 2009;158(2):693–704.

24. Rowe J, Stephan KE, Friston K, Frackowiak R, Lees A, Passingham R. Attention to action in Parkinson’s disease: impaired effective connectivity among frontal cortical regions. Brain. 2002;125(Pt 2):276–289.

25. Rowe JB, Hughes LE, Barker RA, Owen AM. Dynamic causal modelling of effective connectivity from fMRI: are results reproducible and sensitive to Parkinson’s disease and its treatment? Neuroimage. 2010;52(3):1015–1026.

26. Kahan J, Mancini L, Urner M, et al. Therapeutic subthalamic nucleus deep brain stimulation reverses cortico-thalamic coupling during voluntary movements in Parkinson’s disease. PLoS One. 2012;7(12):e50270.

27. Kahan J, Urner M, Moran R, et al. Resting state functional MRI in Parkinson’s disease: the impact of deep brain stimulation on ‘effective’ connectivity. Brain. 2014;137(Pt 4):1130–1144.

28. Bellot E, Coizet V, Warnking J, Moro E, Knoblauch K, Dojat M. Effects of aging on low luminance contrast processing in humans. Neuroimage. 2016;139(October):415–426.

29. Moro E, Bellot E, Meoni S, et al. Visual dysfunction of the superior colliculus in de novo Parkinsonian patients. Annals of neurology. 2020;87:533–546.

30. Postuma RB, Berg D, Stern M, et al. MDS clinical diagnostic criteria for Parkinson’s disease. Mov Disord. 2015;30(12):1591–1601.

31. Hoehn MM, Yahr MD. Parkinsonism: onset, progression and mortality. Neurology. 1967;17(5):427–442.

32. Schneider KA, Kastner S. Visual responses of the human superior colliculus: a high-resolution functional magnetic resonance imaging study. J Neurophysiol. 2005;94(4):2491–2503.

33. Friston KJ, Harrison L, Penny W. Dynamic causal modelling. Neuroimage. 2003;19(4):1273–1302.

34. Penny WD, Stephan KE, Daunizeau J, et al. Comparing families of dynamic causal models. PLoS Comput Biol. 2010;6(3):e1000709.

35. Briggs F, Usrey WM. Corticogeniculate feedback and visual processing in the primate. J Physiol. 2011;589(Pt 1):33–40.

36. Merigan WH, Maunsell JH. How parallel are the primate visual pathway ? Annu Rev Neurosci. 1993;16:369–402.

37. Sherman SM, Koch C. The control of retinogeniculate transmission in the mammalian lateral geniculate nucleus. Exp Brain Res. 1986;63(1):1–20.

38. Benevento LA, Fallon JH. The ascending projections of the superior colliculus in the rhesus monkey (Macaca mulatta). J Comp Neurol. 1975;160(3):339–361.

39. Harting JK, Casagrande VA, Weber JT. The projection of the primate superior colliculus upon the dorsal lateral geniculate nucleus: autoradiographic demonstration of interlaminar distribution of tectogeniculate axons. Brain Res. 1978;150(3):593–599.

40. Munoz DP, Coe BC. Saccade, search and orient--the neural control of saccadic eye movements. Eur J Neurosci. 2011;33(11):1945–1947.

41. Cerkevich CM, Lyon DC, Balaram P, Kaas JH. Distribution of cortical neurons projecting to the superior colliculus in macaque monkeys. Eye Brain. 2014;2014(6):121–137.

42. Künzle H, Akert K. Efferent connections of cortical, area 8 (frontal eye field) in Macaca fascicularis. A reinvestigation using the autoradiographic technique. J Comp Neurol. 1977;173(1):147–164.

43. Kuypers HG, Lawrence DG. Cortical projections to the red nucleus and the brain stem in the Rhesus monkey. Brain Res. 1967;4(2):151–188.

44. Lock TM, Baizer JS, Bender DB. Distribution of corticotectal cells in macaque. Exp Brain Res. 2003;151(4):455–470.

45. Wurtz RH, Goldberg ME. The role of the superior colliculus in visually-evoked eye movements. Bibl Ophthalmol. 1972;82:149–158.

46. Weller RE, Steele GE, Kaas JH. Pulvinar and other subcortical connections of dorsolateral visual cortex in monkeys. J Comp Neurol. 2002;450(3):215–240.

47. Pennartz CMA, Dora S, Muckli L, Lorteije JAM. Towards a unified view on pathways and functions of neural recurrent processing. Trends Neurosci. 2019;42(9):589–603.

48. Georgy L, Celeghin A, Marzi CA, Tamietto M, Ptito A. The superior colliculus is sensitive to gestalt-like stimulus configuration in hemispherectomy patients. Cortex. 2016;81:151–161.

49. Rolland M, Carcenac C, Overton PG, Savasta M, Coizet V. Enhanced visual responses in the superior colliculus and subthalamic nucleus in an animal model of Parkinson’s disease. Neuroscience. 2013;252:277–288.

50. Erskine D, Thomas AJ, Taylor JP, et al. Neuronal loss and alpha-synuclein pathology in the superior colliculus and its relationship to visual hallucinations in Dementia with Lewy Bodies. Am J Geriatr Psychiatry. 2017;25(6):595–604.

51. Frei K, Truong DD. Hallucinations and the spectrum of psychosis in Parkinson’s disease. J Neurol Sci. 2017;374:56–62.

52. Nackaerts E, Michely J, Heremans E, et al. Being on Target: Visual Information during Writing Affects Effective Connectivity in Parkinson’s Disease. Neuroscience. 2018;371:484–494.

53. Goulden N, McKie S, Suckling J, et al. A comparison of permutation and parametric testing for between group effective connectivity differences using DCM. Neuroimage. 2010;50(2):509–515.

54. Daunizeau J, David O, Stephan KE. Dynamic causal modelling: a critical review of the biophysical and statistical foundations. Neuroimage. 2011;58(2):312–322.

55. Stephan KE, Penny WD, Moran RJ, den Ouden HE, Daunizeau J, Friston KJ. Ten simple rules for dynamic causal modeling. Neuroimage. 2010;49(4):3099–3109.

