## Supplementary material for "Effective Connectivity in Subcortical Visual Structures in *De Novo* Patients with Parkinson’s Disease": SM

### Materials and Methods

#### Participants

A detailed visual examination was performed by an ophthalmologist for all participants before the brain MRI. Participants requiring visual correction wore the MediGoggle adult research set (Cambridge Research Systems Ltd, England), interchangeable prescriptive goggles suitable for use in MR environment.

#### fMRI data acquisition

Each fMRI run was composed of four blocks of visual stimulation and five 12-s fixation intervals (fixation cross at the center of the screen), including one between each main block, plus one at the start and one at the end of the run. During the whole duration of each run, each participant was instructed to fixate the fixation cross at the center of the screen and to press a key whenever the orientation of the cross changed. Such an event randomly occurred for the same number of times in each run. This task was used to maintain and control the attention of the subject at the center of the screen. The complete functional session was composed of nine scanning runs, leading to 36 randomized block-presentations of each luminance contrast level.
Experiments were performed using a whole-body 3-Tesla Philips Achieva MRI scanner at the Grenoble MRI facility IRMaGe in France. A 32-channel SENSE head coil was used for image acquisition. For functional scans, a gradient echo planar imaging (EPI MS-FFE) sequence was used (TR/TE = 2000/30 ms, flip angle = 80°, matrix size = 128 x 144, field of view = 192 x 216, 25 transversal slices, slice thickness = 1.5 mm, spatial resolution = 1.5 x 1.5 x 1.5 mm). Brain perfusion measurement was performed in a resting-state condition with a pseudo-continous ASL (pCASL) sequence and the cardiac signal was indirectly recorded using a finger photoplethysmography. These data were used to rule out possible signal contamination by cardiovascular activity and vascular alterations with age or parkinson's disease progression or dopaminergic treatment that could hamper the BOLD signal interpretation.

#### GLM based analysis

Functional data analysis was performed using the single-participant general linear model (GLM) for block-designs with SPM12 (Wellcome Department of Imaging Neuroscience, London, U.K.) implemented in Matlab. For each individual, functional volumes were realigned to correct for head movements and spatially slightly smoothed using a 2-mm FWHM (Full Width at Half Maximum) Gaussian kernel. In our design matrix, movement parameters were entered as nuisance factors. Cardio-respiratory effects are the main cause of source of noise in the BOLD signal in the SC. We introduced the heart rate variability components, computed from the cardiovascular activity measurement, as regressors of non-interest to remove cardio-respiratory components from our fMRI signal. The five conditions of interest (1%, 3%, 5%, 9% and fixation) were modeled as five regressors, constructed as boxcar functions convolved with a canonical hemodynamic response function. We conducted a ROI analysis in the SC, the LGN and the V1, individually defined in each hemisphere based on clusters of activated voxels during all visual conditions versus rest. The V1 was defined as the occipital part activated around the calcarine sulcus. The LGN and the SC were manually refined based on structural information from FGATIR and MPRAGE sequences (see[1] for more details about ROIs definition).

### Time series extraction

Time-series from the ROIs (SC, LGN and V1) were concatenated across runs at the individual level. Contrast images were computed based on the GLM where all conditions were compared to the baseline (fixation). Right and left hemisphere data for each ROI were pooled for each subject. For each participant and each ROI, we extracted the main eigenvariates (adjusted for the participant's effect of interest) from the cluster of activation centered on the peak of activation (uncorrected threshold $p < .001$).

### DCM

DCM considers the brain as a deterministic system where the response in a region of a cortical network is determined by inputs to that region. DCM is a hypothesis-driven technique based on a predefined selection of possible models that are relevant to explore a specific hypothesis. These models describe the possible interactions between ROIs. Inputs to the models to be estimated can be external stimuli (exogenous inputs) or afferences from other regions (endogenous inputs). We used DCM to infer direct connectivity between our ROIs (SC, LGN and V1) based on fMRI time series recorded for these regions. To specify a causal network model in DCM, intrinsic (or endogenous) connections, task-related inputs and possible modulations of these inputs according to the experimental context should be defined. Three sets of parameters are then estimated: the driving input parameters that define how the model responds to external inputs (e.g., visual stimulation; matrix C in DCM), the intrinsic connection parameters (expressed in Hz, matrix A in DCM) that quantify the intrinsic fixed connectivity between the model nodes, i.e. the rate of change in activity in a target region when activity in the source region changes, and the modulatory parameters (expressed in Hz, matrix B in DCM) that define how the effective connectivity may be modulated by experimental conditions such as, in our case, luminance contrast variations. Following models estimation, DCM provides a Bayesian scheme for selecting at the group level, the model of coupling with the highest probability to explain connectivity changes given the observed task-related BOLD responses.[2]

### Model comparison

All 14 models were fed successively with the functional datasets from four groups: controls subjects and untreated PD patients or treated PD patients with dopaminergic treatment (2- or 6-months uptake). They entered a Bayesian Model Selection (BMS) procedure that identified which of the competing models best predicted each dataset. For each model, the model evidence, i.e. the probability of observing the measured data given a specific model, was computed based on the free energy approximation. Model evidence was used by the BMS procedure to rank the models. The exceedance probability (xp) defined the relative superiority that a model was more likely that any other model considered, given the group data. In this context, an exceedance probability of 0.9 for a particular model indicated that this model is 90% more likely to be better than any other model tested given the data. For group-level inference on model structure, we then considered a Random-effects (RFX) BMS, taking into account pathophysiological variations across patients.[3]

### Inference on parameters

Once selected, inference on parameters of the best model was made for each group to assess the significance of the intrinsic connectivity and modulatory parameters (one-sample *t*-test against the null hypothesis, i.e., parameters equal to zero), for more details about DCM methodology, see[4]. Effects of age, disease and medication were analyzed using specific ANOVAs and post-hoc *t*-tests. All statistical analyses were performed using the statistical software Statistica (StatSoft, Europe, Hamburg) for PC version 12.

**Data availability**
All the data are available at Grenoble Institute Neurosciences (Grenoble, France), on request (https://shanoir.irisa.fr/, contact M. Dojat). The data are not publicly available due to privacy restrictions, stated in the document approved by the Institutional Review Board.

### Results

**Healthy Controls.**
**Effect of Age on the intrinsic connectivity**.
Three age-dependent classes were considered: *Young* (n=10, 7 females, 26±3 years), *Middle Age* (n=10, 5 females, 47±4 years) and *Elderly* (n=10, 7 females, 65±3 years). All intrinsic connectivity parameters (expressed in Hz) were positive, suggesting that an increase in activity in each region resulted in an increase in each target region (see Figure 3). A 3x6 analysis of variance (ANOVA) with Age (Young *vs*. Middle age *vs*. Elderly) as between-subjects factor and Connection as within-subjects factor revealed a main effect of Connection ($F(5,135) = 9.06$, $p < 10^{-7}$, $\eta p^2 = 0.25$) but no main effect of Age ($p = 0.61$), and no interaction between Age and Connection ($p = 0.98$). Thus, no age effect was observed whatever the intrinsic connection.
Young: xp = 0.83; Middle age: xp = 0.73; Elderly: xp = 0.70).

**Effect of luminance contrast modulation on the effective connectivity.** For model 10, a 3x4 ANOVA with Age as between-subjects factor and Contrast as within-subjects factor was realized on the modulation parameters. No main effect of Age was observed ($p = 0.47$). However, we found a main effect of Contrast ($F(9,19) = 8.11$, $p < 10^{-4}$, $\eta_p^2 = 0.79$). Interaction between Age and Contrast factors was not significant ($p = 0.28$). To further investigate the main effect of contrast, ANOVAs with Contrast as repeated-measures were realized for each connection and age group. These analyses revealed a modulation by luminance contrast of the SC↔SC connection for the three age groups (Young: $F(3,27) = 9.07$, $p < 10^{-3}$, $\eta_p^2 = 0.50$; Middle age: $F(3,27) = 8.75$, $p < 10^{-3}$, $\eta_p^2 = 0.49$; Elderly: $F(3,27) = 8.99$, $p < 10^{-3}$, $\eta_p^2 = 0.50$;). For the SC→LGN and the SC→V1 connections, a modulation by luminance contrast was revealed for Young (SC→LGN: $F(3,27) = 5.68$, $p < 10^{-2}$, $\eta_p^2 = 0.39$; SC→V1: $F(3,27) = 6.90$, $p < 10^{-2}$, $\eta_p^2 = 0.43$) and Middle age (SC→LGN: $F(3,27) = 11.89$, $p < 10^{-4}$, $\eta_p^2 = 0.57$; SC→V1: $F(3,27) = 4.49$, $p < 10^{-2}$, $\eta_p^2 = 0.33$) groups but not for Elderly subjects (SC→LGN: $p = 0.27$; SC→V1: $p = 0.64$).

### Tables

Table 1. Descriptive statistical values for *de novo* PD patients after 2 and 6 months of dopaminergic treatment and matched controls (mean and standard deviation, SD) for intrinsic connections and modulatory parameters of the best model (model 10 and 6) and the corresponding posterior probabilities. Bold values represent significant values (α = 0.05, two-tailed).

| | **Controls** | | | ***de novo PD patients*** | | | ***PD patients 2m*** | | | ***PD patients 6m*** | | |
|---|---|---|---|---|---|---|---|---|---|---|---|---|
| **Parameters** | **mean** | **SD** | ***p*** | **mean** | **SD** | ***p*** | **mean** | **SD** | ***p*** | **mean** | **SD** | ***p*** |
| ***Intrinsic connections*** | | | | | | | | | | | | |
| LGN→SC | **0.13** | **0.07** | **$< 10^{-10}$** | **0.10** | **0.21** | **0.03** | **0.14** | **0.11** | **$< 10^{-2}$** | 0.10 | 0.20 | 0.17 |
| LGN→V1 | **0.04** | **0.03** | **$< 10^{-6}$** | **0.05** | **0.03** | **$< 10^{-8}$** | 0.002 | 0.05 | 0.91 | **0.08** | **0.09** | **0.02** |
| V1→SC | **0.20** | **0.29** | **$< 10^{-2}$** | **0.24** | **0.14** | **$< 10^{-9}$** | **0.15** | **0.15** | **$< 10^{-2}$** | **0.26** | **0.15** | **$< 10^{-4}$** |
| V1→LGN | **0.09** | **0.08** | **$< 10^{-5}$** | **0.11** | **0.09** | **$< 10^{-6}$** | 0.11 | 0.26 | 0.25 | **0.14** | **0.14** | **$< 10^{-2}$** |
| SC→LGN | **0.06** | **0.06** | **$< 10^{-4}$** | 0.03 | 0.10 | 0.16 | **0.09** | **0.04** | **$< 10^{-5}$** | 0.07 | 0.14 | 0.17 |
| SC→V1 | **0.05** | **0.06** | **$< 10^{-3}$** | **0.08** | **0.06** | **$< 10^{-6}$** | **0.07** | **0.03** | **$< 10^{-5}$** | **0.10** | **0.07** | **$< 10^{-3}$** |
| ***Modulation SC→SC*** | | | | | | | | | | | | |
| 1% | **0.02** | **0.04** | **0.02** | 0.0002 | 0.01 | 0.92 | **0.05** | **0.04** | **$< 10^{-2}$** | / | / | / |
| 3% | **0.04** | **0.04** | **$< 10^{-4}$** | 0.002 | 0.01 | 0.35 | **0.03** | **0.03** | **$< 10^{-2}$** | / | / | / |
| 5% | **0.07** | **0.06** | **$< 10^{-5}$** | 0.003 | 0.01 | 0.16 | **0.02** | **0.02** | **$< 10^{-2}$** | / | / | / |
| 9% | **0.10** | **0.08** | **$< 10^{-6}$** | **0.006** | **0.01** | **$< 10^{-2}$** | 0.03 | 0.09 | 0.36 | / | / | / |
| ***Modulation SC→LGN*** | | | | | | | | | | | | |
| 1% | 0.005 | 0.03 | 0.44 | 0.001 | 0.01 | 0.64 | **0.008** | **0.009** | **0.02** | 0.005 | 0.01 | 0.17 |
| 3% | **0.02** | **0.02** | **$< 10^{-4}$** | 0.003 | 0.01 | 0.35 | 0.003 | 0.01 | 0.41 | 0.006 | 0.01 | 0.11 |
| 5% | **0.02** | **0.02** | **$< 10^{-4}$** | **0.005** | **0.01** | **0.01** | 0.004 | 0.01 | 0.27 | 0.003 | 0.01 | 0.41 |
| 9% | **0.03** | **0.02** | **$< 10^{-7}$** | **0.005** | **0.01** | **0.02** | 0.01 | 0.03 | 0.36 | 0.002 | 0.01 | 0.53 |
| ***Modulation SC→V1*** | | | | | | | | | | | | |
| 1% | 0.001 | 0.005 | 0.35 | 0.002 | 0.04 | 0.81 | **0.004** | **0.004** | **$< 10^{-2}$** | 0.008 | 0.02 | 0.27 |
| 3% | **0.003** | **0.006** | **0.02** | 0.002 | 0.01 | 0.35 | **0.01** | **0.01** | **$< 10^{-2}$** | **0.005** | **0.006** | **0.03** |
| 5% | **0.005** | **0.009** | **0.01** | **0.02** | **0.04** | **0.02** | 0.01 | 0.02 | 0.14 | 0.0001 | 0.005 | 0.95 |
| 9% | **0.01** | **0.02** | **0.02** | **0.04** | **0.03** | **$< 10^{-6}$** | **0.008** | **0.009** | **0.02** | **-0.002** | **0.001** | **$< 10^{-5}$** |
| ***Modulation LGN→SC*** | | | | | | | | | | | | |
| 1% | / | / | / | / | / | / | / | / | / | -0.002 | 0.03 | 0.85 |
| 3% | / | / | / | / | / | / | / | / | / | 0.02 | 0.06 | 0.36 |
| 5% | / | / | / | / | / | / | / | / | / | 0.02 | 0.03 | 0.08 |
| 9% | / | / | / | / | / | / | / | / | / | 0.01 | 0.06 | 0.64 |
| ***Modulation LGN→V1*** | | | | | | | | | | | | |
| 1% | / | / | / | / | / | / | / | / | / | 0.0003 | 0.009 | 0.92 |
| 3% | / | / | / | / | / | / | / | / | / | 0.001 | 0.003 | 0.36 |
| 5% | / | / | / | / | / | / | / | / | / | **0.02** | **0.02** | **$< 10^{-2}$** |
| 9% | / | / | / | / | / | / | / | / | / | **0.01** | **0.01** | **$< 10^{-2}$** |

Table 2. Descriptive statistical values (mean and standard deviation, SD) for endogenous connections and modulatory parameters of the winning model and their posterior probabilities for controls. Bold values represent significant values ($\alpha$ = 0.05, two-tailed).

| | Young | | | Middle age | | | Elderly | | |
|---|---|---|---|---|---|---|---|---|---|
| **Parameters** | **mean** | **SD** | ***p*** | **mean** | **SD** | ***p*** | **mean** | **SD** | ***p*** |
| ***Intrinsic connections*** | | | | | | | | | |
| LGN→SC | **0.13** | **0.14** | **$< 10^{-2}$** | **0.11** | **0.06** | **$< 10^{-4}$** | **0.12** | **0.09** | **$< 10^{-5}$** |
| LGN→V1 | **0.06** | **0.03** | **$< 10^{-5}$** | **0.05** | **0.02** | **$< 10^{-6}$** | **0.04** | **0.03** | **$< 10^{-3}$** |
| V1→SC | **0.26** | **0.12** | **$< 10^{-5}$** | **0.28** | **0.28** | **$< 10^{-2}$** | 0.20 | 0.32 | 0.06 |
| V1→LGN | **0.11** | **0.09** | **$< 10^{-3}$** | **0.13** | **0.09** | **$< 10^{-9}$** | **0.09** | **0.11** | **0.02** |
| SC→LGN | **0.07** | **0.07** | **$< 10^{-2}$** | 0.14 | 0.29 | 0.14 | **0.06** | **0.05** | **$< 10^{-2}$** |
| SC→V1 | **0.07** | **0.04** | **$< 10^{-4}$** | **0.05** | **0.02** | **$< 10^{-5}$** | **0.05** | **0.06** | **0.02** |
| ***Modulation SC→SC*** | | | | | | | | | |
| 1% | **0.05** | **0.04** | **$< 10^{-3}$** | 0.01 | 0.02 | 0.13 | 0.02 | 0.04 | 0.13 |
| 3% | **0.05** | **0.05** | **$< 10^{-2}$** | **0.03** | **0.02** | **$< 10^{-3}$** | **0.05** | **0.03** | **$< 10^{-4}$** |
| 5% | **0.06** | **0.05** | **$< 10^{-2}$** | **0.07** | **0.05** | **$< 10^{-3}$** | **0.06** | **0.05** | **$< 10^{-2}$** |
| 9% | **0.09** | **0.05** | **$< 10^{-4}$** | **0.08** | **0.05** | **$< 10^{-4}$** | **0.08** | **0.07** | **$< 10^{-2}$** |
| ***Modulation SC→LGN*** | | | | | | | | | |
| 1% | **0.02** | **0.01** | **$< 10^{-5}$** | **0.01** | **0.01** | **$< 10^{-2}$** | 0.002 | 0.04 | 0.87 |
| 3% | **0.02** | **0.01** | **$< 10^{-5}$** | **0.03** | **0.02** | **$< 10^{-3}$** | **0.02** | **0.02** | **$< 10^{-2}$** |
| 5% | **0.03** | **0.01** | **$< 10^{-7}$** | **0.04** | **0.01** | **$< 10^{-9}$** | **0.02** | **0.02** | **$< 10^{-2}$** |
| 9% | **0.03** | **0.007** | **$< 10^{-9}$** | **0.04** | **0.01** | **$< 10^{-9}$** | **0.02** | **0.02** | **$< 10^{-2}$** |
| ***Modulation SC→V1*** | | | | | | | | | |
| 1% | **0.005** | **0.006** | **0.02** | 0.003 | 0.01 | 0.35 | 0.002 | 0.04 | 0.87 |
| 3% | **0.008** | **0.007** | **$< 10^{-2}$** | **0.007** | **0.01** | **0.04** | **0.005** | **0.006** | **0.02** |
| 5% | **0.02** | **0.01** | **$< 10^{-5}$** | **0.01** | **0.008** | **$< 10^{-3}$** | 0.008 | 0.02 | 0.17 |
| 9% | **0.02** | **0.01** | **$< 10^{-5}$** | **0.02** | **0.02** | **$< 10^{-2}$** | 0.01 | 0.02 | 0.13 |

### Figures

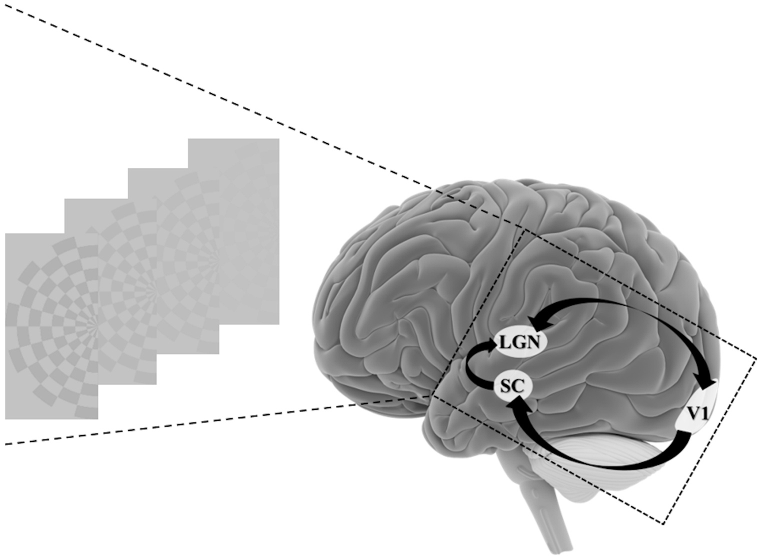


**Figure 1.** The fMRI experience. The visual stimulation that modulates the output of the Superior Colliculus (SC), the Lateral Geniculate Nucleus (LGN) and the first visual area (V1).

`

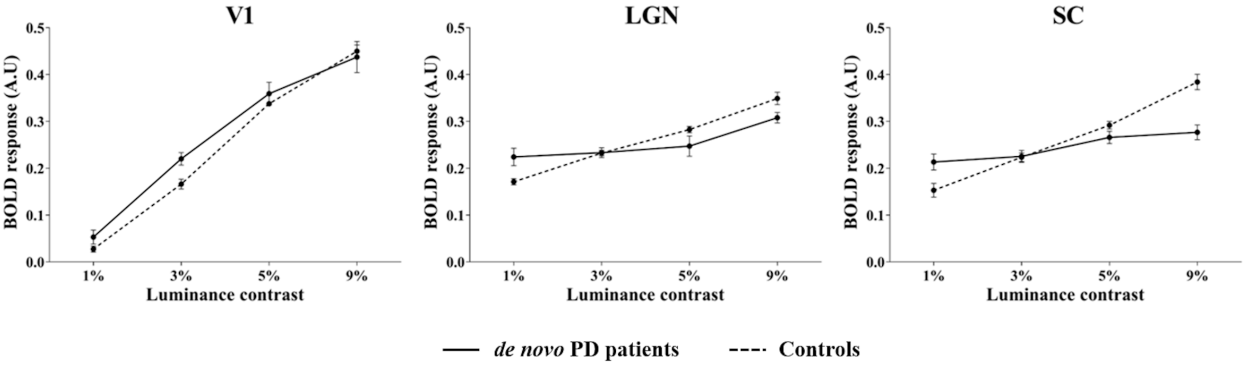


**Figure 2.** Modulation of the BOLD signal following increasing luminance contrast in the first visual area (V1), the Lateral Geniculate Nucleus (LGN) and the Superior Colliculus (SC). Clearly the modulation was hampered in LGN and SC for *de novo* PD patients.

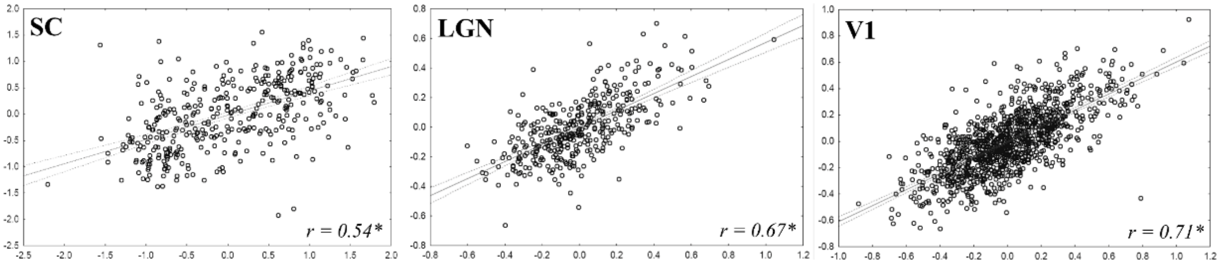


**Figure 3**. Quality of the fitting for our best model (10) between observed and predicted time-series in ROIs (SC, LGN and V1) for a randomly selected individual. Same abbreviations than in Figure 1. * $p < 10^{-51}$.

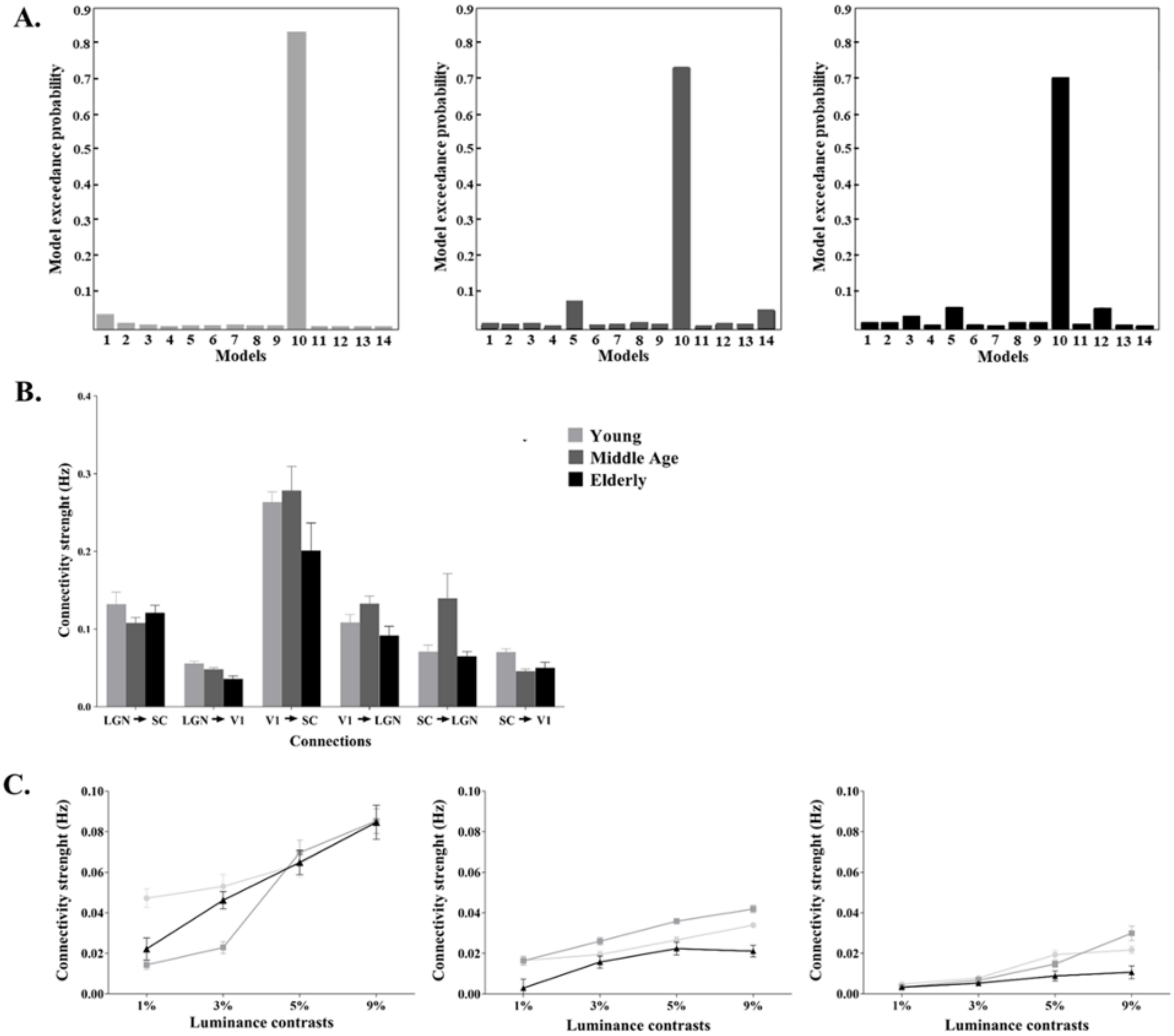


**Figure 4.** Control subjects. A. Exceedance probabilities of the models for the three age groups (left: Young, middle: Middle age, right: Elderly). Model 10 was the best model with random effect Bayesian model selection (RFX) B. Endogeneous connections parameters of the selected model (10) for the three age groups. C. Endogenous connection modulation with luminance contrast (left: self-modulation of SC, middle: modulation of SC to LGN connection, right: modulation of SC to V1 connection). Circle: Young, Square: Middle age, Triangle: Elderly. Endogenous connection and modulatory parameters are expressed in Hz. Same abbreviations than in Figure 1. The vertical bars represent standard deviation.

### References


1. Bellot E, Coizet V, Warnking J, Moro E, Knoblauch K, Dojat M. Effects of aging on low luminance contrast processing in humans. *Neuroimage.* 2016;139(October):415-426.
2. Stephan KE, Penny WD, Daunizeau J, Moran RJ, Friston KJ. Bayesian model selection for group studies. *Neuroimage.* 2009;46(4):1004-1017.
3. Penny WD, Stephan KE, Daunizeau J, et al. Comparing families of dynamic causal models. *PLoS Comput Biol.* 2010;6(3):e1000709.
4. Stephan KE, Penny WD, Moran RJ, den Ouden HE, Daunizeau J, Friston KJ. Ten simple rules for dynamic causal modeling. *Neuroimage.* 2010;49(4):3099-3109.

**Figure 1**

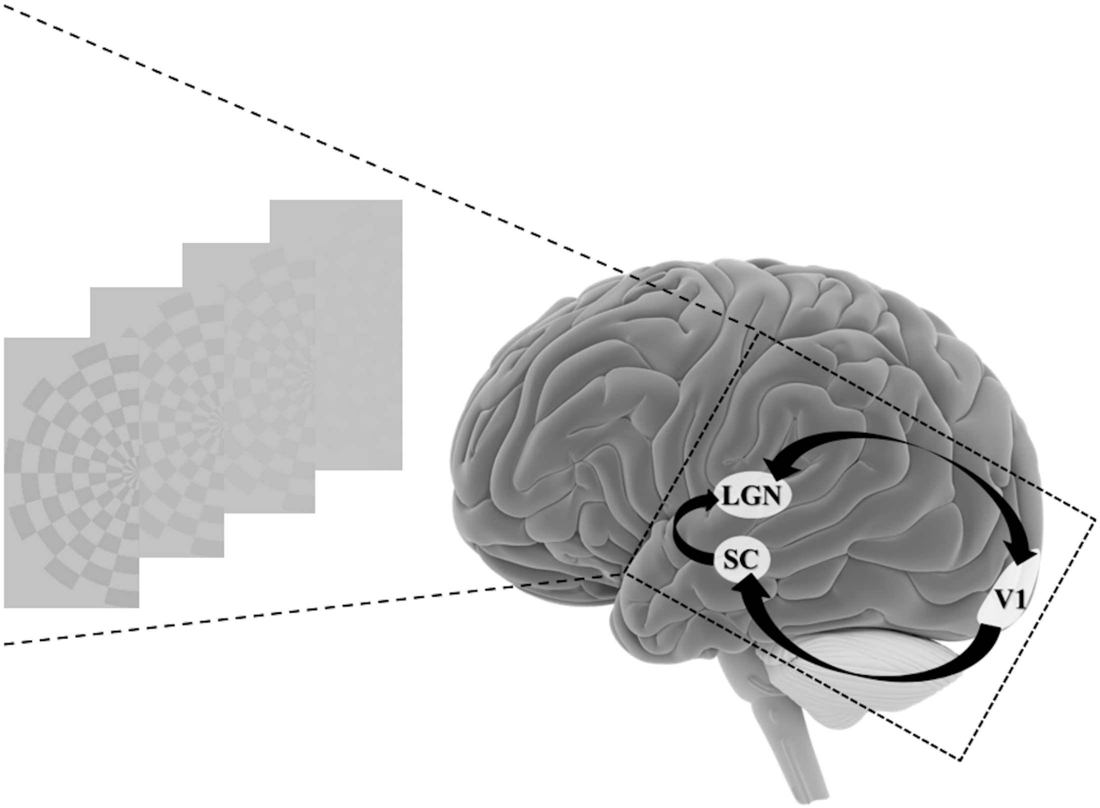


Illustration of the visual stimulus and the three subcortical and cortical regions of interest involved in the first image processing steps and studied with fMRI. Black arrows indicate the anatomical connections based on the literature. SC = superior colliculus, LGN = lateral geniculate nucleus, V1 = primary visual area.

**Figure 2**

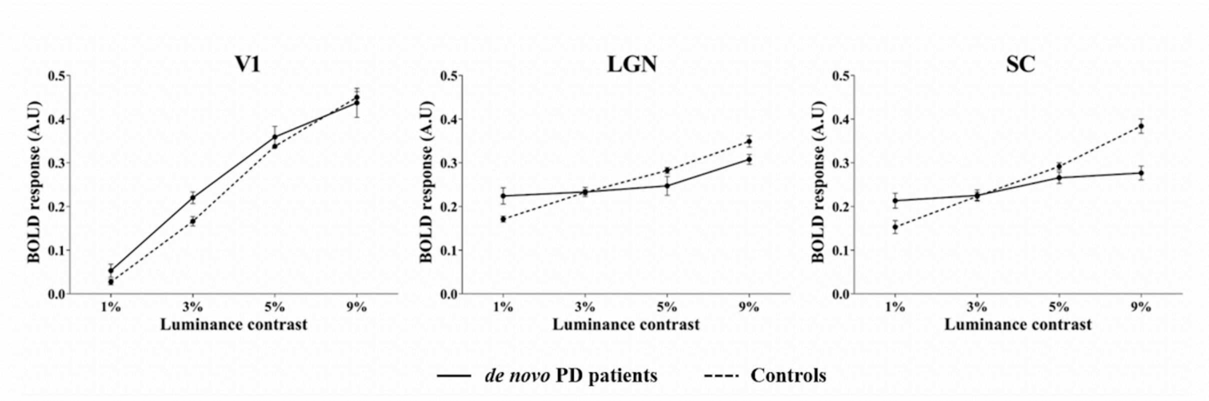


Average variations of the BOLD signal in the three ROIs according to the luminance contrast changes versus fixation for the *de novo* (n=22) Parkinsonian (PD) patients (off medication) and the matched (n=22) controls. Left and right parts of each ROI were combined. Vertical bars indicate standard errors. A.U. = arbitrary unit.

**Figure 3**

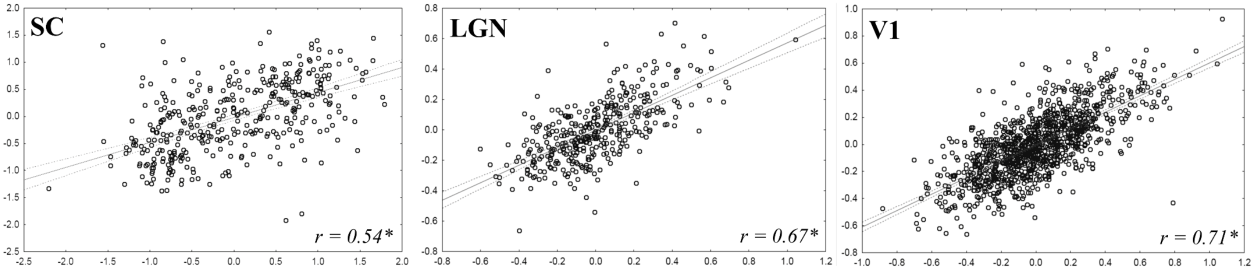


Quality of the fitting for our best model (10) for controls between observed and predicted time-series in ROIs (SC, LGN and V1) for one randomly selected individual. For this individual, we found a correlation coefficient of r = 0.54 for the SC, r = 0.67 for the LGN and r = 0.71 for V1 ($p < 10^{-51}$) between observed and predicted time series.
SC = superior colliculus, LGN = lateral geniculate nucleus, V1 = primary visual area.
* $p < 10^{51}$.

**Figure 4**

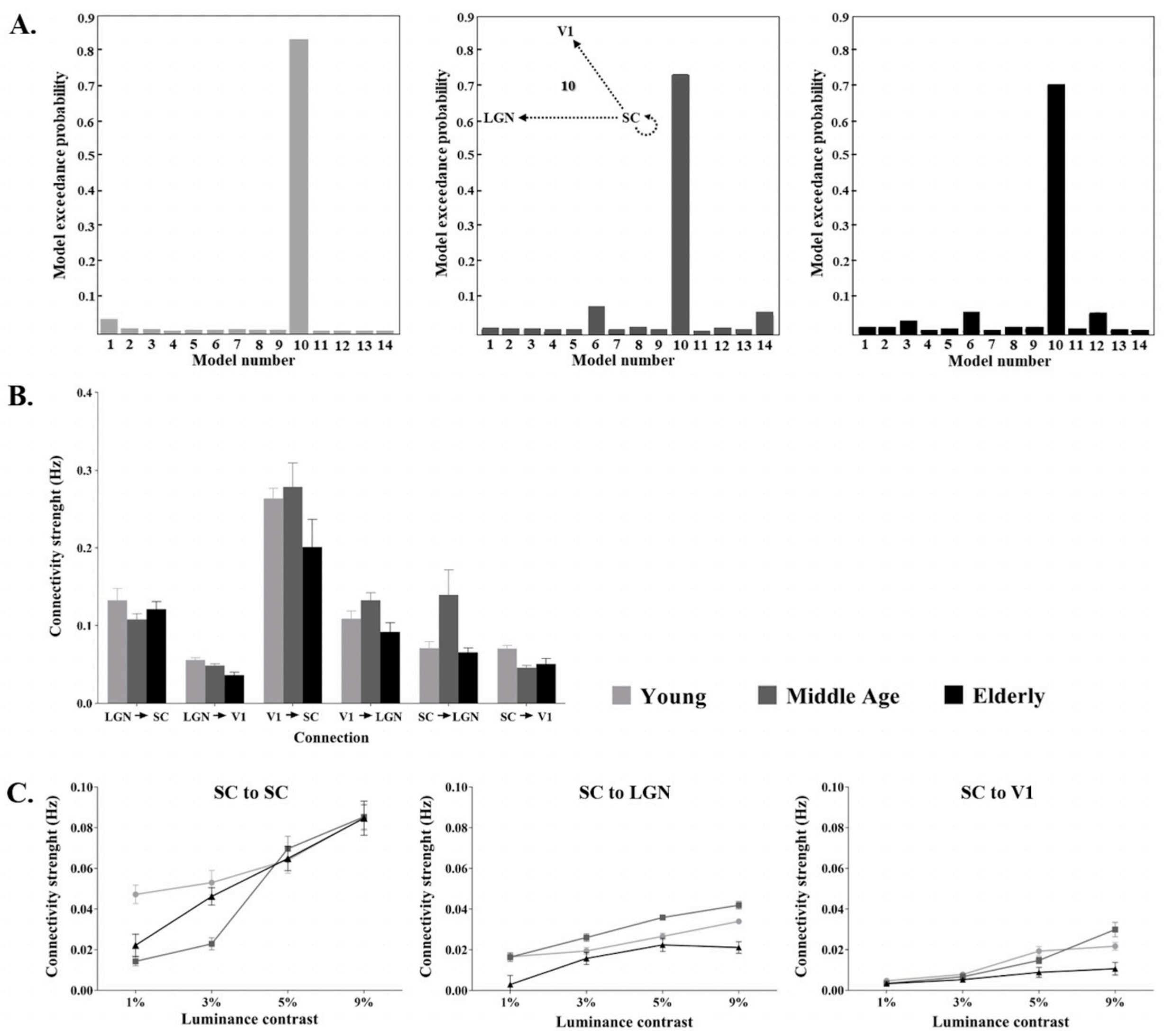


Influence of age on effective connectivity (CT-30). A. Exceedance probabilities of the models for the three age groups of healthy controls (left: Young, middle: Middle age, right: Elderly). For all age ranges, Model 10 (see Insert) was the best model with random effect Bayesian model selection (RFX) B. Strength of the intrinsic connections for the best model for the three age groups. C. Intrinsic connection modulations with luminance contrast. Circle: Young, Square: Middle age, Triangle: Elderly. Intrinsic connection and modulatory parameters are expressed in Hz.
SC = superior colliculus, LGN = lateral geniculate nucleus, V1 = primary visual area. Solid arrows: intrinsic connections. Dashed arrows: connections modulated by luminance. The vertical bars represent standard errors.

Table 1. Descriptive statistical values for matched Controls (CT-22), *de novo* PD patients (PD-dn) and PD patients after 2 and 6 months of dopaminergic treatment (mean and standard deviation, SD) for intrinsic connections and modulatory parameters of the best model (model 10 and 6) and the corresponding posterior probabilities. Bold values represent significant values (α = 0.05, two-tailed).

| | **Controls** | | | ***de novo PD patients*** | | | ***PD patients 2m*** | | | ***PD patients 6m*** | | |
|---|---|---|---|---|---|---|---|---|---|---|---|---|
| **Parameters** | **mean** | **SD** | ***p*** | **mean** | **SD** | ***p*** | **mean** | **SD** | ***p*** | **mean** | **SD** | ***p*** |
| ***Intrinsic connections*** | | | | | | | | | | | | |
| LGN→SC | **0.13** | **0.07** | **$< 10^{-10}$** | **0.10** | **0.21** | **0.03** | **0.14** | **0.11** | **$< 10^{-2}$** | 0.10 | 0.20 | 0.17 |
| LGN→V1 | **0.04** | **0.03** | **$< 10^{-6}$** | **0.05** | **0.03** | **$< 10^{-8}$** | 0.002 | 0.05 | 0.91 | **0.08** | **0.09** | **0.02** |
| V1→SC | **0.20** | **0.29** | **$< 10^{-2}$** | **0.24** | **0.14** | **$< 10^{-9}$** | **0.15** | **0.15** | **$< 10^{-2}$** | **0.26** | **0.15** | **$< 10^{-4}$** |
| V1→LGN | **0.09** | **0.08** | **$< 10^{-5}$** | **0.11** | **0.09** | **$< 10^{-6}$** | 0.11 | 0.26 | 0.25 | **0.14** | **0.14** | **$< 10^{-2}$** |
| SC→LGN | **0.06** | **0.06** | **$< 10^{-4}$** | 0.03 | 0.10 | 0.16 | **0.09** | **0.04** | **$< 10^{-5}$** | 0.07 | 0.14 | 0.17 |
| SC→V1 | **0.05** | **0.06** | **$< 10^{-3}$** | **0.08** | **0.06** | **$< 10^{-6}$** | **0.07** | **0.03** | **$< 10^{-5}$** | **0.10** | **0.07** | **$< 10^{-3}$** |
| ***Modulation SC→SC*** | | | | | | | | | | | | |
| 1% | **0.02** | **0.04** | **0.02** | 0.0002 | 0.01 | 0.92 | **0.05** | **0.04** | **$< 10^{-2}$** | / | / | / |
| 3% | **0.04** | **0.04** | **$< 10^{-4}$** | 0.002 | 0.01 | 0.35 | **0.03** | **0.03** | **$< 10^{-2}$** | / | / | / |
| 5% | **0.07** | **0.06** | **$< 10^{-5}$** | 0.003 | 0.01 | 0.16 | **0.02** | **0.02** | **$< 10^{-2}$** | / | / | / |
| 9% | **0.10** | **0.08** | **$< 10^{-6}$** | **0.006** | **0.01** | **$< 10^{-2}$** | 0.03 | 0.09 | 0.36 | / | / | / |
| ***Modulation SC→LGN*** | | | | | | | | | | | | |
| 1% | 0.005 | 0.03 | 0.44 | 0.001 | 0.01 | 0.64 | **0.008** | **0.009** | **0.02** | 0.005 | 0.01 | 0.17 |
| 3% | **0.02** | **0.02** | **$< 10^{-4}$** | 0.003 | 0.01 | 0.35 | 0.003 | 0.01 | 0.41 | 0.006 | 0.01 | 0.11 |
| 5% | **0.02** | **0.02** | **$< 10^{-4}$** | **0.005** | **0.01** | **0.01** | 0.004 | 0.01 | 0.27 | 0.003 | 0.01 | 0.41 |
| 9% | **0.03** | **0.02** | **$< 10^{-7}$** | **0.005** | **0.01** | **0.02** | 0.01 | 0.03 | 0.36 | 0.002 | 0.01 | 0.53 |
| ***Modulation SC→V1*** | | | | | | | | | | | | |
| 1% | 0.001 | 0.005 | 0.35 | 0.002 | 0.04 | 0.81 | **0.004** | **0.004** | **$< 10^{-2}$** | 0.008 | 0.02 | 0.27 |
| 3% | **0.003** | **0.006** | **0.02** | 0.002 | 0.01 | 0.35 | **0.01** | **0.01** | **$< 10^{-2}$** | **0.005** | **0.006** | **0.03** |
| 5% | **0.005** | **0.009** | **0.01** | **0.02** | **0.04** | **0.02** | 0.01 | 0.02 | 0.14 | 0.0001 | 0.005 | 0.95 |
| 9% | **0.01** | **0.02** | **0.02** | **0.04** | **0.03** | **$< 10^{-6}$** | **0.008** | **0.009** | **0.02** | **-0.002** | **0.001** | **$< 10^{-5}$** |
| ***Modulation LGN→SC*** | | | | | | | | | | | | |
| 1% | / | / | / | / | / | / | / | / | / | -0.002 | 0.03 | 0.85 |
| 3% | / | / | / | / | / | / | / | / | / | 0.02 | 0.06 | 0.36 |
| 5% | / | / | / | / | / | / | / | / | / | 0.02 | 0.03 | 0.08 |
| 9% | / | / | / | / | / | / | / | / | / | 0.01 | 0.06 | 0.64 |
| ***Modulation LGN→V1*** | | | | | | | | | | | | |
| 1% | / | / | / | / | / | / | / | / | / | 0.0003 | 0.009 | 0.92 |
| 3% | / | / | / | / | / | / | / | / | / | 0.001 | 0.003 | 0.36 |
| 5% | / | / | / | / | / | / | / | / | / | **0.02** | **0.02** | **$< 10^{-2}$** |
| 9% | / | / | / | / | / | / | / | / | / | **0.01** | **0.01** | **$< 10^{-2}$** |

Table 2. Descriptive statistical values for controls (CT-30) for three age classes (mean and standard deviation, SD) for intrinsic connections and modulatory parameters of the best model (model 10) and the corresponding posterior probabilities. Bold values represent significant values (α = 0.05, two-tailed).

| | Young | | | Middle age | | | Elderly | | |
|---|---|---|---|---|---|---|---|---|---|
| **Parameters** | **mean** | **SD** | ***p*** | **mean** | **SD** | ***p*** | **mean** | **SD** | ***p*** |
| ***Intrinsic connections*** | | | | | | | | | |
| LGN→SC | **0.13** | **0.14** | **$< 10^{-2}$** | **0.11** | **0.06** | **$< 10^{-4}$** | **0.12** | **0.09** | **$< 10^{-5}$** |
| LGN→V1 | **0.06** | **0.03** | **$< 10^{-5}$** | **0.05** | **0.02** | **$< 10^{-6}$** | **0.04** | **0.03** | **$< 10^{-3}$** |
| V1→SC | **0.26** | **0.12** | **$< 10^{-5}$** | **0.28** | **0.28** | **$< 10^{-2}$** | 0.20 | 0.32 | 0.06 |
| V1→LGN | **0.11** | **0.09** | **$< 10^{-3}$** | **0.13** | **0.09** | **$< 10^{-9}$** | **0.09** | **0.11** | **0.02** |
| SC→LGN | **0.07** | **0.07** | **$< 10^{-2}$** | 0.14 | 0.29 | 0.14 | **0.06** | **0.05** | **$< 10^{-2}$** |
| SC→V1 | **0.07** | **0.04** | **$< 10^{-4}$** | **0.05** | **0.02** | **$< 10^{-5}$** | **0.05** | **0.06** | **0.02** |
| ***Modulation SC→SC*** | | | | | | | | | |
| 1% | **0.05** | **0.04** | **$< 10^{-3}$** | 0.01 | 0.02 | 0.13 | 0.02 | 0.04 | 0.13 |
| 3% | **0.05** | **0.05** | **$< 10^{-2}$** | **0.03** | **0.02** | **$< 10^{-3}$** | **0.05** | **0.03** | **$< 10^{-4}$** |
| 5% | **0.06** | **0.05** | **$< 10^{-2}$** | **0.07** | **0.05** | **$< 10^{-3}$** | **0.06** | **0.05** | **$< 10^{-2}$** |
| 9% | **0.09** | **0.05** | **$< 10^{-4}$** | **0.08** | **0.05** | **$< 10^{-4}$** | **0.08** | **0.07** | **$< 10^{-2}$** |
| ***Modulation SC→LGN*** | | | | | | | | | |
| 1% | **0.02** | **0.01** | **$< 10^{-5}$** | **0.01** | **0.01** | **$< 10^{-2}$** | 0.002 | 0.04 | 0.87 |
| 3% | **0.02** | **0.01** | **$< 10^{-5}$** | **0.03** | **0.02** | **$< 10^{-3}$** | **0.02** | **0.02** | **$< 10^{-2}$** |
| 5% | **0.03** | **0.01** | **$< 10^{-7}$** | **0.04** | **0.01** | **$< 10^{-9}$** | **0.02** | **0.02** | **$< 10^{-2}$** |
| 9% | **0.03** | **0.007** | **$< 10^{-9}$** | **0.04** | **0.01** | **$< 10^{-9}$** | **0.02** | **0.02** | **$< 10^{-2}$** |
| ***Modulation SC→V1*** | | | | | | | | | |
| 1% | **0.005** | **0.006** | **0.02** | 0.003 | 0.01 | 0.35 | 0.002 | 0.04 | 0.87 |
| 3% | **0.008** | **0.007** | **$< 10^{-2}$** | **0.007** | **0.01** | **0.04** | **0.005** | **0.006** | **0.02** |
| 5% | **0.02** | **0.01** | **$< 10^{-5}$** | **0.01** | **0.008** | **$< 10^{-3}$** | 0.008 | 0.02 | 0.17 |
| 9% | **0.02** | **0.01** | **$< 10^{-5}$** | **0.02** | **0.02** | **$< 10^{-2}$** | 0.01 | 0.02 | 0.13 |
